# Most Instability Phases Resolve: Empirical Evidence for Trajectory Plasticity in Multimorbidity Care from Longitudinal Relational Monitoring

**DOI:** 10.64898/2026.04.22.26351537

**Authors:** Carmel M Martin, Keith Stockman, Donald Campbell, Ishbel Henderson

## Abstract

**Background:** The instability plasticity framework proposes that multimorbidity trajectories periodically enter instability phases that are vulnerable to escalation but also potentially modifiable through relational intervention. Whether such phases commonly resolve without acute care, or predominantly progress to hospitalisation, has not been quantified at scale.

**Objective:** To quantify instability window outcomes across a full longitudinal monitoring cohort; to test whether the characteristics distinguishing admitted from resolved windows reflect within-patient trajectory dynamics or between patient severity; and to characterise which patient-reported and operator-rated signals reliably precede admission, using both a curated pilot sub-cohort and the full monitoring cohort with an explicit cross-cohort comparison.

**Methods:** Two complementary analyses were conducted on data from the MonashWatch Patient Journey Record (PaJR) relational telehealth system. Instability windows were identified algorithmically (≥2 consecutive calls with Total_Alerts ≥3) across the full longitudinal dataset (16,383 calls, 244 patients, 2.5 years) and classified by linkage to ED and hospital admission data. Window characteristics were compared at window, patient, and paired within-patient levels. Pre-admission signal cascades were analysed in two configurations: a curated pilot sub-cohort (64 patients, 280 calls, ±10-day window, 103 admissions, December 2016–September 2017) and the full monitoring cohort (175 patients, 1,180 pre-admission calls, ±14-day window, December 2016–July 2019). A three-way cross-cohort comparison decomposed differences between the two configurations into pipeline and population effects.

**Results:** 621 instability windows were identified across 157 patients (64% of the monitored cohort). 67.3% resolved without hospital admission or ED attendance, a rate stable across alert thresholds 1–5. In paired within-patient analysis (n = 70), duration in days (p = 0.002) and multi-domain breadth (p < 0.001) distinguished admitted from resolved windows; alert intensity did not. In the pilot sub-cohort, patient-reported illness prognosis (Q21) was the dominant pre-admission signal (GEE β = +0.058, AUC = 0.647, p–BH = 0.018). This finding did not replicate in the full cohort: Q21 was non-significant (GEE β =− 0.008, p = 0.154, AUC = 0.507). Cross-cohort analysis identified selective curation of the pilot sub-cohort as the primary explanation. In the full cohort, six signals escalated significantly before admission after Benjamini–Hochberg correction: total alerts, health impairment (Q26), red alerts, self-rated health (Q3), patient concerns (Q1), and operator concern (Q34). Health impairment achieved the highest individual AUC (0.605) and showed the longest pre-admission lead. No individual signal exceeded AUC 0.61.

**Conclusions:** Two thirds of instability phases resolve without hospitalisation, providing direct empirical support for trajectory plasticity as a clinically frequent phenomenon. Within the same patient, persistence — in duration and in the consistency of high-severity multi-domain flagging across calls — distinguishes trajectories that tip into admission from those that resolve. The Q21 signal reversal between cohorts illustrates how selective curation can produce compelling but non-replicable findings in monitoring research. In the full population, objective alert signals and operator judgement, rather than patient illness prognosis, carry the pre-admission signal.

## 1. Introduction

Healthcare trajectories among patients with complex multimorbidity are characterised by recurrent cycles of deterioration, acute care utilisation, partial recovery, and renewed instability. For the high-need, high-cost (HNHC) cohort — patients whose care intensity is disproportionately concentrated within health systems [1] — these trajectories are not only intensive but heterogeneous. A recent population-based cohort study of 224,285 HNHC patients in British Columbia identified five distinct cost trajectory groups; standard administrative data could not predict trajectory group membership, suggesting that the mechanisms driving individual trajectory divergence operate at a level invisible to routine health system data [1]. Cross-sectional patient-reported measures improve prediction only modestly [4], consistent with the observation in this cohort that a comprehensive baseline battery — frailty index, resilience, quality of life, and capability — predicted acute admission with a combined ROC of 0.70, a ceiling that static measurement cannot exceed [2].

A companion paper (Martin et al., under review) [3] proposes an instability–plasticity framework grounded in complex adaptive systems theory. Within this framework, multimorbidity trajectories periodically enter instability phases: periods of sustained or clustered signal elevation across care domains that precede acute care transitions. These phases are also intervals of heightened trajectory plasticity, in which relatively small relational interventions may redirect the trajectory before the system tips into hospitalisation. Instability is reconceptualised not only as risk but as opportunity.

Three critical empirical questions follow from this framework. First, what proportion of detected instability phases resolve without hospital admission? If most resolve, this provides direct quantitative evidence that trajectory plasticity is clinically frequent. Second, what distinguishes windows that tip from those that resolve, and do those distinguishing features reflect within-patient trajectory dynamics or between-patient differences in disease burden? Third, which patient-reported and operator-rated signals rise earliest and most reliably before admission? The present paper addresses all three questions through two complementary analyses and an explicit cross-cohort comparison that examines whether the signal profile holds across different cohort configurations and analytical pipelines.

Martin and colleagues previously characterised the ‘pre-hospital syndrome’ in this cohort — alert flags rising significantly in the days before admission [5]. The present analysis extends this work in three ways: quantifying instability window outcomes at scale; using paired within-patient comparisons to control for between-patient confounding; and directly testing whether the pre-admission signal profile is robust to changes in cohort scope and analytical pipeline. The signal reversal identified through cross-cohort comparison is itself reported as a finding about the limits of small-sample monitoring research.

## 2. Materials and Methods

### 2.1. Study Design and Setting

Retrospective observational analysis of longitudinal relational monitoring data from the MonashWatch PaJR telehealth programme within Monash Health, Victoria, Australia. MonashWatch was implemented under the HealthLinks: Chronic Care initiative [6], targeting patients at high predicted risk of recurrent hospitalisation. Enrolled patients received weekly monitoring calls from trained peer health navigators using the PaJR system. Ethics approval was obtained from the Human Research Ethics Committees of Monash Health and Deakin University. Patients were linked across datasets via hashed unit record identifiers; no identifiable data were accessed.

### 2.2. The PaJR Monitoring System

The Patient Journey Record (PaJR) system is a web-based relational monitoring platform supporting structured weekly telephone contacts between peer health navigators and enrolled patients [7,8]. Each call captures patient-reported information across six domains: illness or symptoms, medication, healthcare utilisation, social support, environmental circumstances, and self-care. Within each domain, responses are graded from green (no concern) through amber, continuous, urgent, and immediate red. Total_Alerts represents the count of non-green flags per call across all domains. The six domains parallel the dimensions a general practitioner would systematically assess: symptom burden, medication adherence, health service use, social functioning, living environment, and self-care capacity. Each domain is operationalised through structured branching questions with algorithmic flag generation, supplemented by operator clinical judgement on an overall concern item (Q34).

Self-reported items used in this analysis (Figure 1): Q3 (self-rated health today, 0–5 scale, higher = worse); Q21 (expected health in the next few days; 0 = same or stable, 1 = expects worse, 2 = expects much worse; responses coded 4 in the original dataset represent an active ‘don’t know’ response and were treated as a separate category in sensitivity analyses; primary analyses exclude them; 48% of pre-admission calls have missing Q21, consistent with the branching call structure in which shorter calls do not reach later-sequence questions [7]); Q23 (eating, drinking and toileting function; 1 = problem); Q24 (been outdoors; 1 = not been outdoors); Q25 (sleeping well; 1 = sleep problem); Q26 (health impairment score, derived as 9 minus Q26_Health_score on a 0–9 scale, so higher = worse); Q34 (operator concern; 1 = operator has concerns). Q26 and Q34 were available in the full cohort dataset but not in the curated pilot sub-cohort, and are therefore reported only for ***Table 2***.

**Figure 1.**
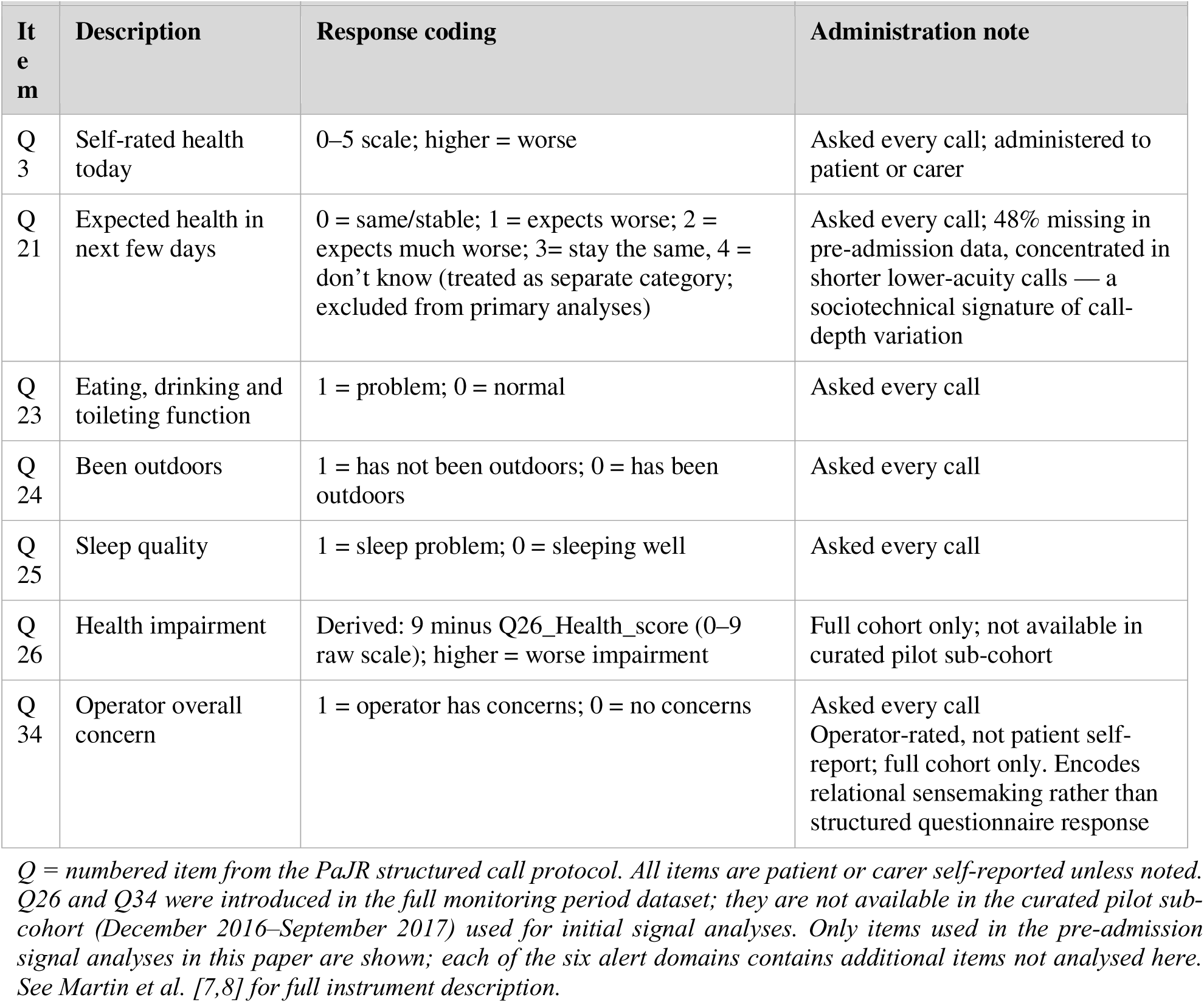
PaJR questionnaire items used in this analysis.

### 2.3. Datasets

Three datasets were used, serving complementary purposes (Table 1). The full longitudinal PaJR dataset comprised 16,383 calls from 244 patients (December 2016 to July 2019), used for instability window detection and outcome classification. Of all calls, 87% were patient-reported and 13% carer-reported; patients contributed a mean of 67.1 calls (SD 40.8) over a mean follow-up of 18.4 months.

**Table 1.** Dataset characteristics.

| Dataset | Purpose | Patients (n) | Records (n) | Period |
| --- | --- | --- | --- | --- |
| Full longitudinal PaJR | Instability window detection and outcome classification | 244 | 16,383 calls | Dec 2016–Jul 2019 |
| Curated pilot sub-cohort | Pre-admission signal cascade (initial analysis) | 64 | 280 calls | Dec 2016–Sep 2017 |
| Full pre/post cohort (v8) | Pre-admission signal cascade (replication and extension) | 175 | 1,180 pre-admission calls | Dec 2016–Jul 2019 |
| ED linkage | Outcome classification and admission route verification | 243 | 2,219 ED records | Jul 2016–Jul 2019 |

*The curated pilot sub-cohort and full pre/post cohort (v8) are explicitly compared in Section 3.4 to decompose cross-cohort differences.*

**Table 2.** Instability window characteristics by outcome.

| Characteristic | Resolved<br>(n=380) | ED-only<br>(n=38) | Admitted<br>(n=197) | p (R vs A) |
| --- | --- | --- | --- | --- |
| Duration (calls), median [IQR] | 2.0 [2.0–3.0] | 2.0 [2.0–4.0] | 3.0 [2.0–4.0] | p=0.001** |
| Duration (days), median [IQR] | 8.0 [7.0–15.2] | 13.0 [7.0–21.0] | 14.0 [7.0–22.0] | p<0.001*** |
| Mean alerts/call, median [IQR] | 5.0 [4.0–6.5] | 5.3 [4.5–7.3] | 5.7 [4.5–7.0] | p=0.005** |
| Peak alerts, median [IQR] | 7.0 [5.0–9.0] | 7.5 [5.0–10.0] | 8.0 [5.0–10.0] | p=0.001** |
| Multi-domain index (red alerts), median [IQR] | 0.5 [0.3–1.0] | 0.9 [0.5–1.0] | 1.0 [0.5–1.1] | p<0.001*** |
| Multi-domain index (any alerts), median [IQR] | 2.0 [1.7–2.5] | 2.5 [2.0–3.0] | 2.5 [2.0–2.8] | p<0.001*** |

*Window-level Mann–Whitney tests. R = resolved; A = admitted. Multi-domain index (red alerts) = mean number of domains with $\geq 1$ Urgent or Immediate Red flag per call. Multi-domain index (any alerts) = mean number of domains with $\geq 1$ Amber or Red flag per call. \*\*\* p<0.001; \*\* p<0.01.*

The curated pilot pre/post admission sub-cohort comprised 280 calls from 64 patients within a ±10-day observation period around 103 acute emergency admissions occurring between December 2016 and September 2017. This sub-cohort was manually curated from the original pilot phase of the monitoring programme. Structured investigation subsequently identified that the curation included 103 of 199 eligible admissions during this period; no consistent selection rule was identified, and the inclusion rate was negatively correlated with total admission count per patient. This selection is described in detail in Section 3.4 as it bears directly on the interpretation of the Q21 findings.

The full pre/post admission dataset was constructed from the complete monitoring period (December 2016 to July 2019) using a corrected analytical pipeline. Each monitoring call was linked to its nearest emergency hospital admission occurring within ±14 days, with nearest-admission de-duplication applied (each call maps to exactly one admission). This produced 2,025 call–admission pairs across 175 patients with at least one emergency admission; 1,180 of these were pre-admission calls.

### 2.4. Instability Window Detection

An instability window was defined as a sequence of ≥2 consecutive monitoring calls in which Total_Alerts ≥3. This threshold captures meaningful multi-signal elevation across domains, while excluding isolated single-domain fluctuations. Duration is measured in monitoring calls (the natural unit of the detection algorithm). Approximate calendar duration assumes a median inter-call interval of 7 days in this dataset (range 1–247 days); calendar duration approximations should be interpreted accordingly. Window duration distributions are right-skewed; medians are reported throughout. Sensitivity analysis confirmed stability of findings across alert thresholds 1–5 (Table 6).

Each window was linked to ED and hospital admission data via hashed patient identifiers, classifying outcomes within 14 days of the final alert call: resolved (no ED attendance or admission), ED-only (ED attendance without inpatient admission; Short Stay Observation Unit classified as ED-only), admitted (departure to inpatient ward, ICU, CCU, or medical assessment and planning unit), or ongoing (window still open at end of monitoring record). Inpatient admission was verified against ED departure status fields; 96.6% of admissions were matched to an ED presentation occurring a median 4.7 hours earlier, confirming the linkage.

### 2.5. Within-Patient Analysis

To distinguish within-patient trajectory dynamics from between-patient severity differences, window characteristics were compared at three levels: (a) window-level Mann–Whitney tests treating all windows as independent; (b) patient-level comparisons aggregating each patient’s windows into a single mean value per outcome category; and (c) paired within-patient Wilcoxon signed-rank tests restricted to the 70 patients who contributed both admitted and resolved windows. The paired analysis is the most interpretively sound comparison, eliminating between-patient confounding entirely. The window-level analysis is presented for completeness but requires caution in interpretation as it conflates within-patient and between-patient variation.

### 2.6. Pre-Admission Signal Cascade Analysis

For each cohort configuration, pre-admission calls were partitioned into non-overlapping time bands relative to the admission event. Three complementary statistical approaches were applied to each signal: (a) Spearman rank correlation of the signal value with DaysFromAdmission (positive rho = signal higher when closer to admission); (b) Mann–Whitney U comparison of ‘imminent’ (within 7 days) versus ‘earlier’ (more than 7 days) pre-admission calls; and (c) Generalised Estimating Equations (GEE) linear models with exchangeable within-patient correlation structure, accounting for the clustering of repeated calls within patients. GEE is the primary and most defensible test; Spearman and Mann–Whitney results are presented to allow assessment of robustness. All p-values were adjusted using the Benjamini–Hochberg false discovery rate method applied within each analytical family. Logistic GEE models were used to derive AUC values for the capacity of individual signals to classify calls within 7 days of admission.

A prospective lead-lag analysis examined whether activity-domain signals at call N predicted Q21-worse responses at call N+1 (within 14 days), using Fisher’s exact test. This prospective design is more demanding than concurrent association tests and its results are highlighted accordingly.

### 2.7. Cross-Cohort Comparison

A three-way cross-cohort comparison was conducted: (i) the original curated pilot sub-cohort (v3); (ii) the same admissions re-analysed with the corrected pipeline and extended ±14-day window (v7-pilot); and (iii) the full monitoring cohort with the corrected pipeline (v8-full). Comparing v3 with v7-pilot isolates pipeline and window effects for the same admissions. Comparing v7-pilot with v8-full isolates the effect of broadening from a curated subset to the full monitored population. Patient identity and data consistency were verified by cross-checking hashed identifiers and signal values across files.

### 2.8. Statistical Analysis

Descriptive statistics are reported as medians [IQR] for skewed distributions and means (SD) for approximately normal distributions. The analysis was conducted in R using the geepack package for GEE models and base R for other tests. Analysis scripts (analysis_paper_2_v8.R) are available from the corresponding author. The study is observational; findings should be interpreted as hypothesis-generating rather than confirmatory.

## 3. Results

### 3.1. Study Population

244 patients were enrolled in MonashWatch between December 2016 and August 2017; no new enrolments occurred thereafter. The cohort was closed, with attrition throughout the monitoring period as the programme wound down. Mean follow-up was 18.4 months (SD 8.7); 217 patients were active at peak programme size (July 2017), declining to 77 at the data extract date (July 2019). Mean calls per patient were 67.1 (SD 40.8, median 68 [IQR 37–92]). 87.2% of calls were patient-reported and 12.8% carer-reported. Alert signals showed a highly skewed distribution: mean 1.46 alerts per call (SD 2.47, median 0, maximum 21); 53.7% of calls had no alerts. Red alerts (Urgent and Immediate Red flags only) were present in 17.2% of calls, mean 0.26 per call.

### 3.2. Instability Window Detection and Outcomes

Applying the detection algorithm (Total_Alerts ≥3, ≥2 consecutive calls) to the full longitudinal dataset identified 621 instability windows across 157 patients (64% of the monitored cohort). The mean number of windows per patient with at least one window was 3.96 (SD 3.44, range 1–16), equivalent to a median rate of 2.2 windows per patient per year of follow-up. Window duration was right-skewed (median 10 days [IQR 7–21], mean 19.8 days, maximum 471 days).

Outcome classification yielded: 380 windows resolved without acute care (61.2%), 38 ED-only (6.1%), 197 inpatient admitted (31.7%), and 6 ongoing at end of monitoring (1.0%). The combined non-admission rate was 67.3% (418 of 621 windows). Sensitivity analysis across alert thresholds confirmed this finding: the non-admission rate ranged from 71.1% at threshold ≥1 to 65.8% at threshold ≥5, with the primary threshold (≥3) at 67.3% (Table 6).

#### 3.2.1. Within-Patient Analysis: Separating Trajectory Dynamics from Severity

Window-level comparisons conflate within-patient trajectory dynamics with between-patient differences in disease burden. The three-level analysis (Table 4) addresses this by progressively controlling for between-patient confounding.

**Table 4.** Three-level comparison of window characteristics by outcome (paired within-patient analysis, n=70).

| Characteristic | Median difference (admitted – resolved) | Paired p |
| --- | --- | --- |
| Duration (calls) | 0.0 | $p=0.070\sim$ |
| Duration (days) | +3.6 days | $p=0.002^{**}$ |
| Mean alerts/call | +0.19 | $p=0.124$ ns |
| Peak alerts | +0.67 | $p=0.057\sim$ |
| Multi-domain index (red alerts) | +0.18 | $p<0.001^{***}$ |
| Multi-domain index (any alerts) | +0.15 | $p=0.004^{**}$ |

*Wilcoxon signed-rank test, $n=70$ patients with both admitted and resolved windows. Difference = patient mean (admitted windows) minus patient mean (resolved windows). $\sim p<0.10$ ; $^{**} p<0.01$ ; $^{***} p<0.001$ ; ns = not significant.*

At the patient level (aggregating each patient’s windows), only duration in days (p=0.003) and the multi-domain indices (red alerts p<0.001; any alerts p<0.001) remained significant; alert intensity did not (mean alerts p=0.11, peak alerts p=0.06). The paired within-patient comparison (n=70 patients with both admitted and resolved windows) is the most interpretively sound analysis. Within the same patient, admitted windows lasted a median of 3.6 days longer (p=0.002) and showed greater multi-domain breadth (red-alert index median difference +0.18, p<0.001; any-alert index median difference +0.15, p=0.004). Alert intensity did not significantly distinguish admitted from resolved windows generated by the same patient (mean alerts p=0.12, peak alerts p=0.06).

This finding has direct implications for monitoring system design. Alert intensity in absolute terms reflects between-patient differences in disease burden, not within-patient trajectory dynamics approaching a tipping point. What distinguishes trajectories that tip, within the same patient, is whether the instability persists over a longer period and whether high-severity flagging is sustained across calls rather than coming and going. The temporal-persistence dimension of the multi-domain index — how consistently any domain is flagged at high severity across the calls of a window — is the discriminating signal.

### 3.3. Pre-Admission Signal Cascade: Curated Pilot Sub-Cohort

This section reports results from the original curated pilot sub-cohort (64 patients, 280 calls, ±10-day observation window, 103 admissions, December 2016–September 2017). Findings should be read as initial observations from a pilot dataset. Section 3.4 reports the full cohort results and Section 3.5 provides the cross-cohort comparison.

Total alerts and red alerts were elevated throughout the pre-admission period and spiked acutely in the final 24 hours (Total_Alerts mean 6.0 [SD 4.6]; red alerts 1.75 [SD 1.16]). Self-rated health (Q3) showed no significant pre-admission trend (Spearman rho = +0.039, p = 0.62).

Patient-reported illness prognosis (Q21) showed the strongest association with proximity to admission (Spearman rho = +0.303, p = 0.006; p–BH = 0.042). The proportion of calls in which patients expected to feel worse increased progressively from 25.8% at 10–7 days before admission to 75.0% in the final 24 hours (Table 5). GEE models confirmed Q21 as significant after clustering and BH correction (β = +0.058, SE = 0.019, p = 0.003, p–BH = 0.018; n = 80 observations from 40 patient clusters after excluding ‘don’t know’ and non-response). Q23 (eating, drinking, toileting) was borderline significant (GEE p = 0.018, p–BH = 0.049). GEE logistic regression confirmed Q21 as the strongest single predictor of calls within 7 days of admission (AUC = 0.647).

**Table 5.** Self-reported signal values by time interval relative to admission — curated pilot sub-cohort (64 patients).

| Interval | n | Q3 mean (SD) | Q21 mean (SD) | Q21> 0 % | Alerts mean (SD) | Red alerts mean (SD) | Sleep % | Outdoors % |
| --- | --- | --- | --- | --- | --- | --- | --- | --- |
| –10 to –7d | 53 | 2.70 (0.91) | 0.26 (0.44) | 25.8% | 3.38 (3.39) | 0.70 (0.99) | 44.1% | 21.9% |
| –7 to –3d | 69 | 2.42 (1.02) | 0.50 (0.58) | 46.4% | 2.20 (2.95) | 0.49 (0.92) | 29.0% | 27.3% |
| –3 to –1d | 30 | 2.93 (1.08) | 0.71 (0.69) | 58.8% | 4.93 (5.09) | 1.20 (1.52) | 30.0% | 25.0% |
| –1 to 0d | 8 | 3.29 (1.60) | 1.00 (0.82) | 75.0% | 6.00 (4.60) | 1.75 (1.16) | 33.3% | 60.0% |
| 0–3d after | 15 | 3.20 (1.08) | 0.45 (0.69) | 36.4% | 5.60 (3.89) | 2.13 (1.46) | 30.8% | 66.7% |

| Interval | n | Q3 mean (SD) | Q21 mean (SD) | Q21>0 % | Alerts mean (SD) | Red alerts mean (SD) | Sleep % | Outdoors % |
| --- | --- | --- | --- | --- | --- | --- | --- | --- |
| 3–7d after | 54 | 2.54 (1.00) | 0.32 (0.47) | 32.4% | 3.39 (3.73) | 0.98 (1.00) | 16.2% | 41.0% |
| 7–10d after | 51 | 2.55 (0.92) | 0.44 (0.58) | 40.7% | 3.08 (3.17) | 0.63 (0.87) | 44.8% | 44.8% |

*Q3: self-rated health (0–5, higher = worse). Q21: illness prognosis (0 = stable, 1 = worse, 2 = much worse); means and proportions computed on non-missing subset excluding ‘don’t know’ and non-response. n = total calls in interval.*

### 3.4. Pre-Admission Signal Cascade: Full Monitoring Cohort

This section reports results from the full monitoring cohort (175 patients, 1,180 pre-admission calls, ±14-day window, corrected pipeline).

The alert cascade was pronounced: mean total alerts increased from 1.73 at 14–10 days before admission to 4.47 on the day before admission, while red alerts increased 4.6-fold from 0.29 to 1.33 over the same period. Self-rated health deteriorated in parallel (mean Q3 from 2.17 to 3.10). After admission, signals partially recovered without returning to the pre-admission outer-band level (total alerts 2.38 at 10–14 days post; red alerts 0.52), consistent with the clinical picture of patients returning to an elevated baseline of chronic complexity.

After Benjamini–Hochberg correction, six signals were significant in GEE models (Table below): total alerts (β = +0.125, p–BH < 0.001), health impairment Q26 (β = +0.085, p–BH < 0.001, AUC = 0.605), red alerts (β = +0.035, p–BH < 0.001), self-rated health Q3 (β = +0.035, p–BH < 0.001), patient concerns Q1 (β = +0.017, p–BH < 0.001), and operator concern Q34 (β = +0.014, p–BH < 0.001). Q23 was additionally significant (β = +0.010, p–BH = 0.014). Patient-reported illness prognosis (Q21) was non-significant (β =− 0.008, p = 0.154, AUC = 0.507). Q24, Q25, and Q21 in all coding variants were non-significant.

Health impairment (Q26) is notable for two features. It achieved the highest individual AUC (0.605) of any signal. Uniquely, its GEE beta strengthened when the analysis window was extended from ±10 to ±14 days (the only signal to do so), suggesting elevation beginning as early as 14 days before admission — a longer pre-admission lead than other signals. Operator concern (Q34) was the most consistent signal across both the pilot subcohort and the full cohort, and its status as a strong predictor reflects the operator’s synthesis of today’s structured responses against their accumulated longitudinal knowledge of the patient.

The lead-lag analysis (1,461 consecutive call pairs, mean gap 6.6 days) identified Q23 as the only signal with a prospective association: patients who flagged eating, drinking, or toileting problems at call N were more likely to report expected worsening (Q21 > 0) at call N+1 (OR = 1.86, Fisher p = 0.033, n = 387 complete pairs). The same-call association between Q23 and Q21 was non-significant, confirming the prospective rather than concurrent nature of this relationship. Q24 and Q25 were non-significant in the lead-lag analysis.

No individual signal or combination achieved an AUC above 0.61. The best performing combination in the sub-cohort (SRH + Q21, AUC = 0.594) did not generalise to the full cohort. This AUC ceiling reflects the limitations of single-call point-in-time analysis, not necessarily the clinical utility of the monitoring programme, which operates through longitudinal trajectory interpretation by operators and clinicians with accumulated patient knowledge.

Future analysis of operator narrative notes and free-text comments accompanying each monitoring call is currently underway and may substantially extend the findings reported here. Operator narratives may illuminate the relational and contextual information that Q34 as a binary concern flag approximates but cannot fully represent: the specific language patterns, expressions of uncertainty, references to trajectory deviation from a patient’s norm, and qualitative clinical observations that operators record alongside the structured questionnaire responses. This analysis may also shed light on the Q21 “don’t know” finding — the response category carrying the highest prospective admission rate (11.7%) — by examining whether prognostic uncertainty as expressed in narrative language is itself a detectable risk marker. Findings from the narrative analysis will be incorporated into a follow-up paper.

### 3.5. Cross-Cohort Comparison: The Q21 Signal Reversal

The signal profiles diverge substantially across cohort configurations; Table 7 summarises the key results.

**Table 6.**
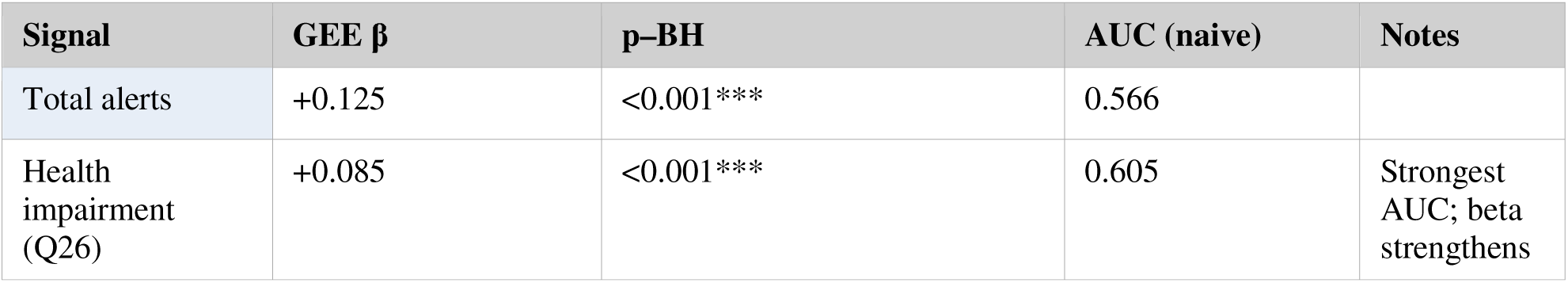
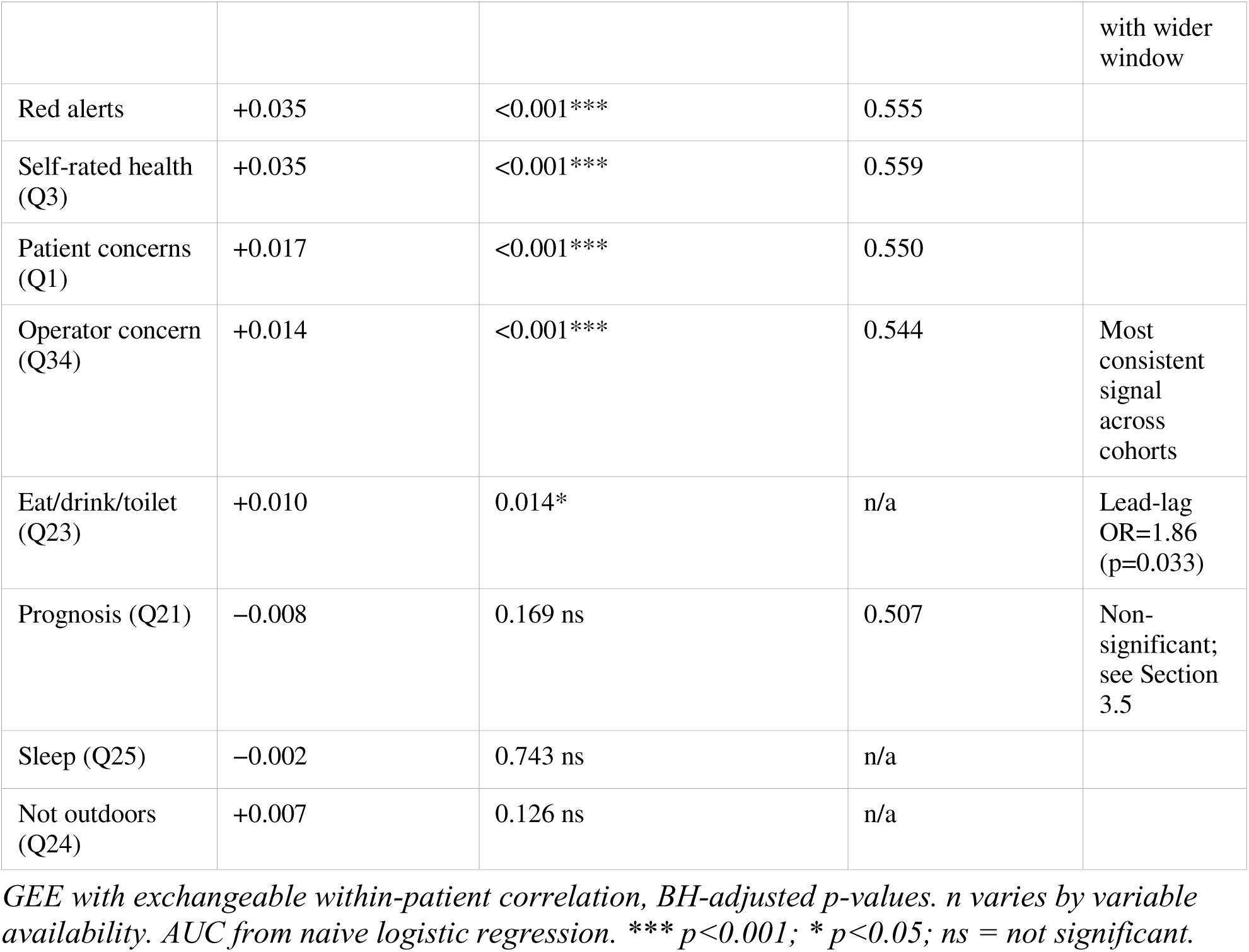
(inset). Pre-admission signal results — full cohort (GEE, n=1,180 calls, 175 patients).

**Table 7.** Cross-cohort comparison of key signals.

| Signal | v3 curated pilot | v7-pilot (corrected pipeline) | v8-full cohort |
| --- | --- | --- | --- |
| Q21 GEE $\beta$ (p-BH) | primary: +0.058 (0.018*) | attenuated: +0.035 (0.043*) | non-sig: -0.008 (0.169) |
| Q21 AUC | 0.647 | 0.563 | 0.507 |
| Total alerts GEE $\beta$ (p) | ns (0.108) | significant (0.002**) | primary: +0.125 (<0.001***) |
| Red alerts GEE $\beta$ (p) | n/a | significant (<0.001***) | significant: +0.035 (<0.001***) |
| Self-rated health (Q3) (p) | ns (0.207) | significant (0.0001***) | significant: +0.035 (<0.001***) |
| Operator concern (Q34) (p) | not available | significant (<0.001***) | significant: +0.014 (<0.001***) |
| Q23 (p) | sig (0.018*) | ns (0.39) | sig (0.009**) |

*v3 = original curated pilot sub-cohort ( $\pm 10$ -day window, 64 patients, 103 admissions). v7-pilot = same admissions, corrected pipeline ( $\pm 14$ -day window). v8-full = full monitoring cohort, corrected pipeline ( $\pm 14$ -day window, 175 patients). \* $p < 0.05$ ; \*\* $p < 0.01$ ; \*\*\* $p < 0.001$ ; ns = not significant.*

Two effects explain the Q21 reversal. First, pipeline effects (v3 → v7-pilot, same admissions): the wider ±14-day window adds lower-acuity outer-band calls that dilute the Q21 escalation slope, attenuating the beta by approximately 40%. This is a methodological window-expansion effect. Second, population broadening effects (v7-pilot → v8-full): the full cohort has a chronically elevated Q21 baseline (approximately 55–65% of calls have Q21 > 0 across all pre-admission windows), leaving no room for the escalation that was visible against the pilot sub-cohort’s 26% baseline.

Structured investigation of the curation confirmed that the pilot sub-cohort included 103 of 199 eligible admissions (52%) during this period with no consistent selection rule. The inclusion rate was negatively correlated with the total number of admissions per patient: patients with fewer admissions were more likely to be included. The same patients, when examined across all their admissions in the full cohort, show a 59.2% Q21>0 rate — compared to 42.5% in the curated subset. The curation selected, without a systematic rule, specific admission episodes where Q21 happened to discriminate most compellingly. This is a curation artefact, not a replicable biological signal.

The alert signal profile shows the reverse pattern: total and red alerts were non-significant in v3 (p = 0.108 and marginally significant respectively) and became increasingly significant and consistent as cohort scope broadened. Operator concern (Q34) was not available in the original v3 dataset but was strongly significant in both v7-pilot and v8-full, making it the most robust signal in the analysis.

**Table 8.**
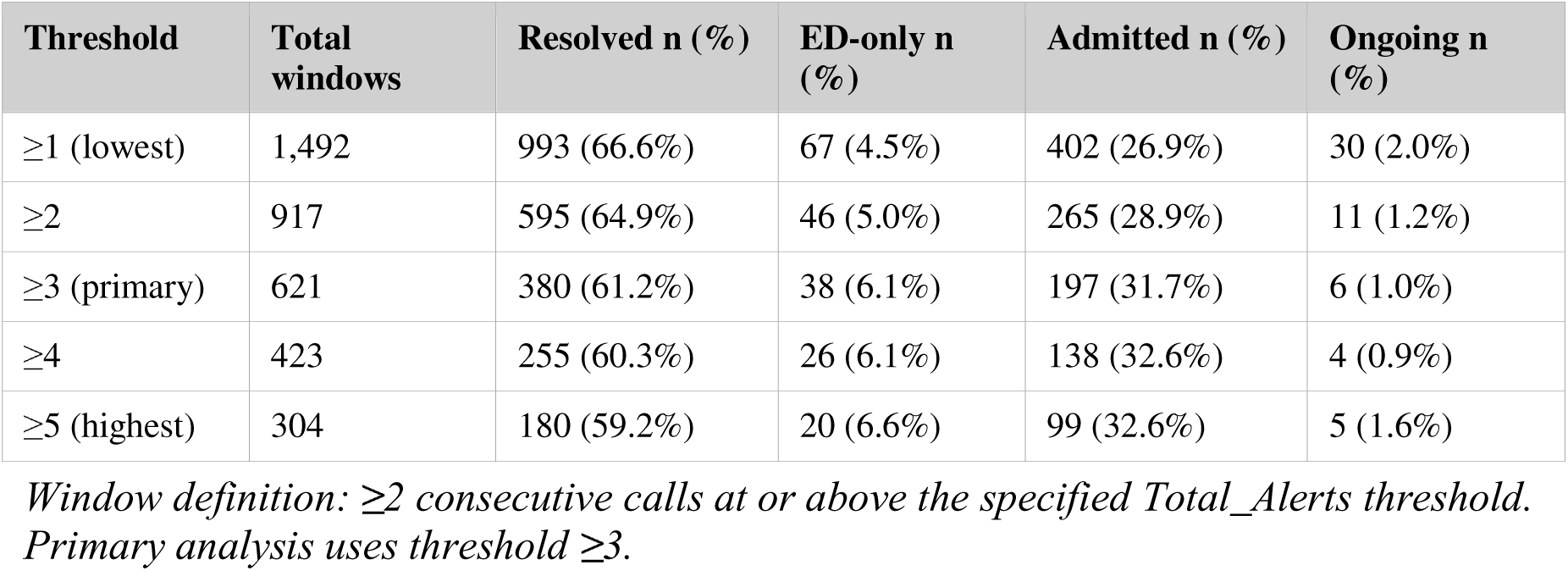
Instability window outcome sensitivity analysis across alert thresholds.

## 4. Discussion

### 4.1. Trajectory Plasticity as an Empirical Finding

The central finding of this study is that 67% of detected instability windows resolved without hospital admission, a rate stable across alert thresholds 1–5 and across patient subgroups. This is the empirical foundation of trajectory plasticity as a clinically frequent, not merely theoretical, phenomenon. Within a high-risk population already enrolled in monitoring, individual trajectories regularly enter and exit instability phases, and most do so without hospitalisation. This reframes how monitoring systems should be understood: they are not primarily admission prediction tools but observation platforms for the natural ebb and flow of multimorbidity, with the majority of detected episodes representing opportunities for clinical awareness rather than inevitable precursors to acute care.

### 4.2. What Distinguishes Windows That Tip

The within-patient paired analysis (n = 70) provides the most interpretively sound answer to what distinguishes trajectories that tip into admission from those that resolve. Within the same patient, the discriminating features are persistence in time (longer window duration) and the consistency of high-severity multi-domain flagging across calls. Alert intensity — the absolute number of flags per call — does not discriminate once between-patient severity is controlled. This finding is methodologically important: window-level intensity differences reflect between-patient confounding (sicker patients generate higher alert counts overall and also experience more admissions), not within-patient trajectory dynamics approaching a tipping point.

The persistence interpretation has a resilience-theory reading. When high-severity flagging is intermittent across the calls of a window, the patient and their support system appear to be recovering between calls and the trajectory is self-correcting. When high-severity flagging is sustained from call to call, the compensatory capacity that absorbs intermittent disturbances is no longer keeping pace. This is consistent with the complex adaptive systems framing in which resilience is an emergent property of the whole system [9]: the temporal pattern of severe alerting, not the instantaneous level, signals whether resilience is holding. In this dataset, the discriminating feature is whether high severity persists across calls, not whether it spreads across domains in any single call — although the multi-domain index reflects both dimensions because it averages across calls within the window.

The practical implication is direct. Monitoring systems designed around instantaneous alert thresholds miss the dynamics that distinguish trajectories approaching a tipping point from those that are transiently unstable. Trajectory surveillance should track: the running duration in days of any open instability window; the count of consecutive calls within an open window in which the multi-domain index at high severity is greater than zero; and the trend across calls rather than the value at any single call.

### 4.3. The Pre-Admission Signal Profile in the Full Cohort

In the full monitoring cohort, the pre-admission signal is carried by objective alert measures and operator judgement rather than patient illness prognosis. Total alerts more than doubled, red alerts increased 4.6-fold, and self-rated health, patient concerns, and operator concern all escalated significantly in GEE models accounting for within-patient clustering. Health impairment (Q26) achieved the highest individual discriminative performance (AUC = 0.605) and the longest pre-admission lead, with its beta strengthening rather than attenuating when the analysis window was extended to 14 days. No individual signal or combination exceeded AUC 0.61, confirming that per-call discrimination is modest regardless of which signal is used.

Operator concern (Q34) deserves particular attention. It was the most consistent signal across both the pilot sub-cohort and the full cohort, and its robustness is not coincidental. Q34 encodes the output of a relational sensemaking process — the operator’s synthesis of today’s call against their accumulated knowledge of this patient’s baseline, typical fluctuations, and previous concerns. This is precisely the information that single-call AUC analysis cannot capture. The finding that the most consistent quantitative signal is the one that most directly reflects human relational knowledge suggests that the analytical approach (per-call discrimination from structured responses) underestimates the clinical utility of the monitoring programme, which operates through longitudinal trajectory interpretation rather than single-call scoring.

These findings position PaJR as a sociotechnical system rather than a measurement instrument. In sociotechnical systems theory, technical components (structured questionnaires, algorithmic alert rules, flag thresholds) and social components (the relational encounter, operator sensemaking, accumulated patient knowledge, post-call clinician debrief) are mutually constitutive: neither layer produces the clinical value independently [15]. The Q34 finding is the empirical expression of this: the most consistent pre-admission signal in the dataset is the one that most directly encodes the social layer — operator judgement formed through repeated relational contact — rather than any measure generated by the technical layer alone. The AUC ceiling of 0.61 reflects not a failure of measurement but a consequence of analysing the technical layer in isolation from the social layer within which it operates. Notably, the 2012 Irish PaJR implementation — which used the full sociotechnical system including narrative text, machine learning over conversational data, and care guide clinical judgement — achieved 100% sensitivity for hospital admissions [7]. The gap between that result and the AUC of 0.61 obtained here from structured items alone is the empirical measure of what the social layer contributes. Q21’s selective completion pattern (48% missingness, concentrated in shorter lower-acuity calls) is a further sociotechnical signature: whether the navigator reached the question depended on call length and conversational depth — properties of the relational encounter — rather than on instrument design. The original PaJR implementation reported that 50% of calls lasted 2–3 minutes (‘no problem’ calls) and only 25% exceeded 5 minutes [7]; Q21, positioned later in the call sequence, was simply not reached in the shorter calls.

The Q23 lead-lag finding (OR = 1.86, p = 0.033) is noteworthy because it is prospective: Q23 problems at call N predicted Q21-worse responses at call N+1, with no concurrent association. This temporal ordering is what a monitoring programme needs. The GEE significance of Q23 was unstable across sub-cohorts, but its prospective lead-lag result is more demanding and its consistency supports treating it as a candidate early-warning signal worthy of further investigation.

### 4.4. Signal replication across cohort configurations: a methodological case study

The reversal of the prognosis signal between cohorts — from a statistically significant finding in the curated pilot sub-cohort to a non-significant result in the full population — is an important methodological finding in its own right. It illustrates precisely how selective curation of pilot data can produce apparently robust results that do not survive replication at scale.

The curation selected 103 of 199 eligible admissions (52%) with no consistent rule, biasing toward episodes where patients had a low Q21 baseline that escalated dramatically toward admission. The same patients, when all their admissions are included, show a chronically elevated Q21 baseline with no room for such escalation. This is not a failure of the prognosis question as a clinical concept: asking patients whether they expect to feel worse is sensible and the signal may well carry prognostic information in individual cases. The failure was methodological: the pilot data selected the episodes where the signal happened to discriminate most compellingly and then reported those results as if they were characteristic of the monitored population.

This has broader implications for monitoring research. Monitoring datasets are complex, longitudinal, and heterogeneous. Small pilot cohorts, particularly those assembled with any degree of manual curation, carry substantial risk of selection artefacts that will not survive population-level replication. Three-way cross-cohort comparisons of the kind conducted here — same admissions with corrected pipeline, then full population — provide a discipline that few analyses achieve but that should be standard practice before any monitoring signal finding is presented as established.

### 4.5. Implications for Trajectory Stewardship

These findings support shifting monitoring system design from instantaneous alert-level monitoring toward trajectory-persistence surveillance. The clinical implication is that a monitoring system should track, for each patient, the running duration of any open instability window and the temporal consistency of high-severity flagging across calls — not just whether today’s call generated alerts. When a window has been in the alert state for two or more weeks without resolution, the patient’s internal redirection capacity may be exhausted and the trajectory is at its most sensitive to clinical response.

Operator concern and health impairment are the signals that most reliably anticipate the pre-admission period in the full population. Monitoring system design should support and amplify operator judgement rather than attempting to replace it with automated scoring. The Q34 finding suggests the relational dimensions of the monitoring encounter — the operator knowing the patient, recognising deviation from their individual baseline, and being positioned to act on it — carry clinical information that structured questionnaire responses cannot fully capture.

Sociotechnical system design principles suggest that improving programme effectiveness requires investing in both layers jointly: refining the alert algorithm and questionnaire coverage on the technical side, while simultaneously strengthening the relational infrastructure — operator training, continuity of patient assignment, post-call debrief protocols, and clinician integration — through which the technical signals are interpreted and acted upon. Optimising the technical layer alone, without attending to the social layer that gives it meaning, is unlikely to improve clinical outcomes.

### 4.6. Limitations

The instability window detection algorithm is a pragmatic analytical construct. The primary threshold and minimum-length parameters were chosen a priori; sensitivity analysis provides reassurance but not external validation. Window outcomes are classified by proximity to ED and admission data within a 14-day symmetric window; some windows classified as admitted may include calls occurring after discharge.

Patients with only admitted windows had substantially shorter monitoring follow-up than those with only resolved windows (25% vs 6% with less than 6 months). The paired within-patient analysis controls for this bias; unpaired comparisons should be interpreted in this context. The cohort is from a single health service in Victoria, Australia; generalisability to other settings is not established.

The Q21 GEE significance was sensitive to window width and cohort composition in ways described in detail in Section 3.5. Q21 analyses in the pilot sub-cohort are based on 80 observations from 40 patient clusters after excluding ‘don’t know’ responses; the sample size constrains power and precision. The ‘don’t know’ response category (Q21 value = 4) had the highest prospective admission rate of any Q21 category (11.7%) and may itself carry prognostic information; its exclusion from primary analyses is an analytical decision that warrants future investigation.

The observational design cannot distinguish between instability windows that resolved through relational monitoring, through other community or clinical care, or spontaneously. The monitoring system creates the opportunity; establishing the causal contribution of the monitoring itself to resolution requires a comparison-group design with broader care event linkage.

## 5. Conclusion

Two thirds of instability phases in this monitored HNHC multimorbidity cohort resolved without hospitalisation, providing direct empirical support for trajectory plasticity as a clinically frequent phenomenon. Within the same patient, persistence — in time and in the consistency of high-severity multi-domain flagging across calls — distinguishes trajectories that tip into admission from those that resolve. Alert intensity does not discriminate once between-patient severity is controlled.

The signal profile that reliably precedes admission in the full population is carried by objective alert signals and operator concern, not by patient illness prognosis. The Q21 signal that appeared compelling in the curated pilot sub-cohort did not replicate in the full cohort; cross-cohort analysis identified selective curation as the primary explanation, providing a case study in the methodological risks of small-sample monitoring research.

These findings suggest that monitoring system design should prioritise tracking trajectory persistence and multi-domain spread over time, rather than instantaneous alert thresholds, and should preserve and support the relational operator judgement that is the most consistent pre-admission signal Taken together, the Q34 finding, the AUC ceiling, and the pattern of Q21 selective completion converge on a single interpretive frame: PaJR functions as a sociotechnical system, in which the structured questionnaire and alert algorithm constitute a technical layer whose clinical value is realised through the social layer of relational operator knowledge, longitudinal patient familiarity, and post-call clinical sensemaking. Evaluation frameworks that assess per-call signal discrimination alone will systematically underestimate the programme’s utility and misidentify where improvement investment should be directed.

identified in this analysis.

## Acknowledgements

The authors acknowledge the telecare guides, health coaches, and patients of the MonashWatch programme and the peer health navigators whose sustained engagement generated the longitudinal dataset on which this study depends.

## Author Contributions

Conceptualisation: C.M.M.; methodology: C.M.M. and K.S.; formal analysis: K.S.; data curation and data analysis: K.S. and D.C.; writing—original draft: C.M.M.; writing—review and editing: K.S., D.C., I.H. All authors have read and agreed to the submitted version.

## Data Availability Statement

The datasets analysed in this study are not publicly available due to ethics and privacy restrictions governing the MonashWatch programme. De-identified summary data and analysis code (analysis_paper_2_v8.R) are available from the corresponding author on reasonable request.

